# AMPLITUDE PERFORMANCE SUBTYPES IN PARKINSON’S DISEASE

**DOI:** 10.64898/2026.07.08.26357552

**Authors:** Antje S. Mefferd, Kris Tjaden, Mary S. Dietrich, Amy E. Brown

## Abstract

**Purpose:** The purpose of this study was to identify subgroups of talkers with Parkinson’s disease (PD) with shared tongue, lip, and jaw articulatory amplitude behaviors. The study also sought to identify demographic and clinical features that can distinguish the identified kinematic subgroups.

**Methods:** 53 talkers with PD and 54 controls participated. Articulatory amplitudes of the tongue, lip, and jaw were measured during a paragraph reading task using three-dimensional electromagnetic articulography. Amplitude performance profiles of the tongue, lip, and jaw were established for each talker with PD by referencing their performance to that of controls. These profiles were submitted to a hierarchical cluster analysis to identify kinematic-based subgroups. Amplitude performances were compared across subgroups to determine between-group patterns. Demographic and clinical features (e.g., age, sex, disease duration, selected perceptual speech characteristics, dysarthria severity) were compared across the identified kinematic subgroups.

**Results:** Four main kinematic subgroups with differing amplitude performance profiles were identified. One subgroup exhibited normal to mildly exaggerated or mildly reduced amplitudes and was labeled “preclinical subgroup” (*n* = 16). Three subgroups exhibited pronounced amplitude reductions of either the tongue (*n* = 10), the tongue and lips (*n* = 12), or the tongue, lips, and jaw (*n* = 10). In addition, there were five talkers with PD whose performance profiles did not align with the identified four subgroups. Their performance was characterized by either pronounced amplitude exaggerations or mildly reduced jaw and lip amplitudes and exaggerated tongue amplitudes. None of the demographic or clinical features differed significantly between the main four subgroups.

**Conclusion:** Findings suggest that the extent to which hypokinesia manifests within the articulatory subsystem can vary in talkers with PD. Longitudinal studies are needed to determine if these subgroups represent different stages of disease progression or distinctly different manifestations of the disease within the articulatory subsystem.

## INTRODUCTION

PD is a common neurodegenerative disease that affects approximately 12 million people worldwide (Luo et al., 2025). It is associated with primary motor symptoms such as bradykinesia and hypokinesia, rigidity, tremor, and gait instability and secondary motor symptoms such as micrographia, shuffled gait, and dysarthria (e.g., Moustafa et al., 2016). Individuals with PD can also experience non-motor symptoms such as cognitive impairment, dysautonomia, and sleep disorders (e.g., Schapira, Chaudhuri, & Jenner, 2017). However, the extent to which motor and non-motor symptoms manifest in each person can vary greatly.

Over the last decade, research efforts have been increasingly directed towards the identification of subtypes of PD to aid research studies that seek to delineate the underlying pathophysiology of the disease and improve our understanding of the disease progression. The identification of subtypes can also improve the design of clinical trials and pave the way to more personalized medicine (e.g., Deng et al., 2025; Mestre et al., 2021). Traditionally, PD has been associated with two motor subtypes based on gross motor symptoms: the tremor predominant and the akinetic-rigid subtype (Zetusky, Jankovic, & Pirozzolo, 1985). Based on the Unified Parkinson’s Disease Rating Scale (UPDRS) motor subtypes of PD have also been divided into a tremor-dominant, a mixed/indeterminate, and a postural instability gait difficulty subtype (Jankovic et al., 1990). However, more recently, non-motor subtypes and genetic subtypes have also been established (e.g., Deng et al., 2022; Dulski et al., 2015; Sauerbier et al., 2016).

Although motor subtypes of PD are differentiated based on gross motor symptoms, speech is a motor skill that is also affected in individuals with PD. In fact, approximately 90% of individuals with PD develop dysarthria as part of their disease progression (Ho et al., 1998; Logemann et al., 1978). The dysarthria type that is typically associated with PD is hypokinetic dysarthria. Although these talkers can vary greatly in their dysarthria symptoms (e.g., breathy, hoarse, or harsh vocal quality and fast, variable, or slow speech rate; Duffy, 2019), these variable speech characteristics do not seem to map onto the specific motor subtypes of PD (Brown & Spencer, 2020).

Within the speech motor system, hypokinesia and rigidity are thought to underlie dysarthria symptoms (e.g., Hanson et al., 1984; Kearney et al., 2017; Ma, Lau, & Thyagarajan, 2020; Mefferd et al., 2026, Walsh & Smith, 2012; Yunusova et al., 2008). For example, imprecise speech, a symptom of articulatory subsystem impairment, has been associated with reduced articulatory amplitudes (Thompson & Kim, 2025). However, so far, speech kinematic studies that directly examined articulatory movements of talkers with PD have yielded mixed results. That is, some studies did not observe reduced movement amplitudes (e.g., Ackermann et al., 1997; Konczak et al., 2004; Mefferd & Dietrich, 2019) while others observed reduced movement amplitudes in only a subset of the articulators (e.g., Connor et al., 1998; Kearney et al., 2017; Yunusova et al., 2008). By contrast, some studies reported reduced movement amplitudes across all articulators (e.g., Kuruvilla-Dugdale & Mefferd, 2022). In other studies, tongue movements were found to be even larger in talkers with PD than in controls (Thies et al., 2023; Wong et al., 2011).

It is possible that mixed findings of kinematic studies were due to methodological variables such as the use of different speech stimuli or different measurement approaches. However, the lack of convergence on amplitude differences between talkers with PD and controls may also be due to a high level of variability in articulatory performance across talkers with PD (e.g., Ackermann et al., 1997; Yunusova et al., 2008). To address this issue, we have previously identified subgroups of talkers with PD using auditory-perceptual assessments of articulatory performance (Mefferd & Dietrich, 2019; Mefferd et al., 2026). That is, speech language pathologists listened to a speech sample of each study participant and determined by consensus ratings the presence (PDdys+artic) or absence (PDdys-artic) of an articulatory subsystem impairment. Mefferd and Dietrich (2019) showed that talkers in the PDdys-artic subgroup produced larger jaw movements than controls during the production of the diphthong /ai/. However, this finding was not replicated in Mefferd et al., (2026), which included a larger sample of talkers with PD and examined articulatory movements during a paragraph reading task. However, the relatively high variability of the jaw performance within the PDdys-artic subgroup in Mefferd et al. (2026) suggests that some of the talkers in this subgroup exhibited abnormally large jaw movements while others did not. Thus, grouping talkers with PD merely based on perceptually defined speech subsystem impairment profile may not be ideal to identify subgroups of talkers with similar articulatory performance patterns.

The current study builds upon our prior work and sought to identify kinematic subgroups of talkers with PD based on their articulatory performance profiles using a data-driven cluster analysis approach. A better understanding of potential differences in articulatory performance profiles of the tongue, lip, and jaw in talkers with PD is important for several reasons. For example, such information is critically needed to delineate how articulator-specific amplitude deficits contribute to a talker’s speech intelligibility loss. Based on such insights, it will be possible to precisely pinpoint the articulator(s) needing to improve or be compensated for by less affected articulators. Relatedly, knowledge about potentially different articulatory performance profiles of talkers with PD can help to better understand the articulatory mechanisms that underlie variable response patterns to therapeutic speech interventions. It is well-known that some talkers with PD show poor responses to behavioral speech interventions such as clear, loud, or slow speech cues (Kim et al., 2026). It has also been shown that clear, loud, and slow speech cues elicit articulator-specific amplitude changes. For example, slow speech has been shown to facilitate predominantly independent tongue movements while loud and clear speech engage the jaw and tongue (Mefferd, 2017; Mefferd & Dietrich, 2019). Thus, a better understanding of a talker’s articulatory performance profile may allow clinicians to select a speech intervention that is known to specifically target a talker’s performance deficits.

However, it is not currently feasible for clinicians to examine articulatory performance directly using movement tracking devices such as the electromagnetic articulograph. Therefore, a secondary goal of this study was to explore if kinematic-based subgroups of talkers with PD were associated with demographic and/or clinical features such as age, sex, age of disease onset, disease duration, treatment type (i.e., medication only versus deep brain stimulation), dysarthria severity, intelligibility, or specific perceptual speech features (e.g., consonant imprecision, fast rate).

Demographic features such as age and sex may predict specific kinematic subgroups in PD since these factors have been shown to impact articulatory behavior of healthy controls (e.g., Hermes et al., 2018; Simpson, 2001). Relatedly, the age of disease onset may be associated with specific articulatory behaviors as an earlier disease onset may result in different articulatory impairment patterns than a later disease onset due to overall age affecting the articulatory behavior. In addition, clinical features such as the disease duration and dysarthria severity may be predictive of the articulatory patterns as they may be associated with various disease stages (i.e., early versus late). Intelligibility may also be differentially affected by different articulatory performance patterns as amplitude reduction of the jaw or lips, for example, may not be as detrimental to intelligibility as amplitude reduction of the tongue. Furthermore, there is emerging evidence that deep brain stimulation (DBS) may affect articulatory behavior of talkers with PD (e.g., Nip et al., 2023). Therefore, it is possible that the treatment type (medication versus DBS) may be associated with specific kinematic subtypes of PD. Finally, perceptual speech features can vary greatly across talkers with PD. Particularly rate-related perceptual speech features (i.e., overall articulatory rate, increased rate in segments, short rushes of speech), as well as loudness-related perceptual speech features (i.e., overall loudness, vocal decay) may be associated with specific kinematic subtypes as kinematic studies have shown robust rate and loudness effects on articulatory amplitudes in healthy talkers and talkers with PD (e.g., Mefferd & Green, 2010; Mefferd, 2015). In addition, reduced stress, a perceptual speech feature that arises from insufficient adjustments in duration, pitch, and vocal intensity (e.g., Lehiste, 1970), is a potential perceptual feature that may be associated with a specific kinematic subtype of PD due to known rate and loudness effects on articulatory performance. Therefore, the purpose of the current study was to 1) identify kinematic subtypes in talkers with PD and 2) determine if demographic and/or clinical features are associated with specific kinematic subgroups of PD.

## METHODS

The current study included 53 talkers with PD (29 males, 24 females) and 54 controls (29 males, 25 females). This dataset is the same as the one used by Mefferd et al. (2026) with the addition of 13 talkers with PD and 14 controls. In fact, the current study is a secondary analysis of the kinematic data reported in Mefferd et al. with the inclusion of additional talkers with PD and controls. The current study is focused on the variable performance profiles of talkers with PD while Mefferd et al. reported group findings of talkers with PD and controls. The study was approved by the Vanderbilt University Medical Center institutional review board (IRB#150655) and all participants provided written consent prior to data collection. All participants were recruited as part of a larger, ongoing research project and were compensated for their time and effort.

All participants spoke American English as their native language, had no history of a neurological condition other than PD, did not report any history of a speech, language, or hearing impairment, and passed a cognitive screening (the Mini Mental Status Examination, Folstein & Folstein, 2012) with a score of at least 20 out of 30 points to ensure that they could follow the instructions during the experiment and complete the experimental tasks. One control did not complete the cognitive screening due to time constraints but reported no cognitive difficulties.

All participants with PD were diagnosed by a board-certified neurologist and were taking anti-Parkinsonian medication. They all completed the data collection within two hours of their last medication dose. Five participants with PD had undergone DBS as part of their treatment of the disease and had their device turned on during the data collection. 16 participants with PD had been enrolled in speech therapy to address their dysarthria at some point prior to the data collection and ten participants with PD were enrolled in speech therapy at the time of data collection.

All participants completed the Sentence Intelligibility Test (SIT, Yorkston et al., 2007) to document their intelligibility level. For each SIT test, a new set of eleven sentences was generated. Three research assistants orthographically transcribed each SIT recording, and the three scores were then averaged for each participant. For 12 controls, the mean SIT score was only based on two transcribed SIT scores. Four participants (two controls, two talkers with PD) had missing SIT recordings due to an experimenter error. The SIT recording was also used to determine the severity of the dysarthria of each talker with PD. Two speech language pathologists with expertise in motor speech disorders (one being the first author) independently rated the dysarthria severity of each talker with PD using a 5-point Likert scale (0 = normal, 1 = mild, 2 = moderate, 3 = severe, 4 = profound). The speech language pathologists then compared their ratings, discussed any discrepant ratings, and provided a final consensus rating for each talker with PD. Table 1 presents the demographic information of all participants along with their SIT scores, the dysarthria severity level, and information on the disease duration of talkers with PD. As can be seen, talkers with PD were generally highly intelligible. The median dysarthria severity rating indicated a moderate dysarthria.

**Table 1.**
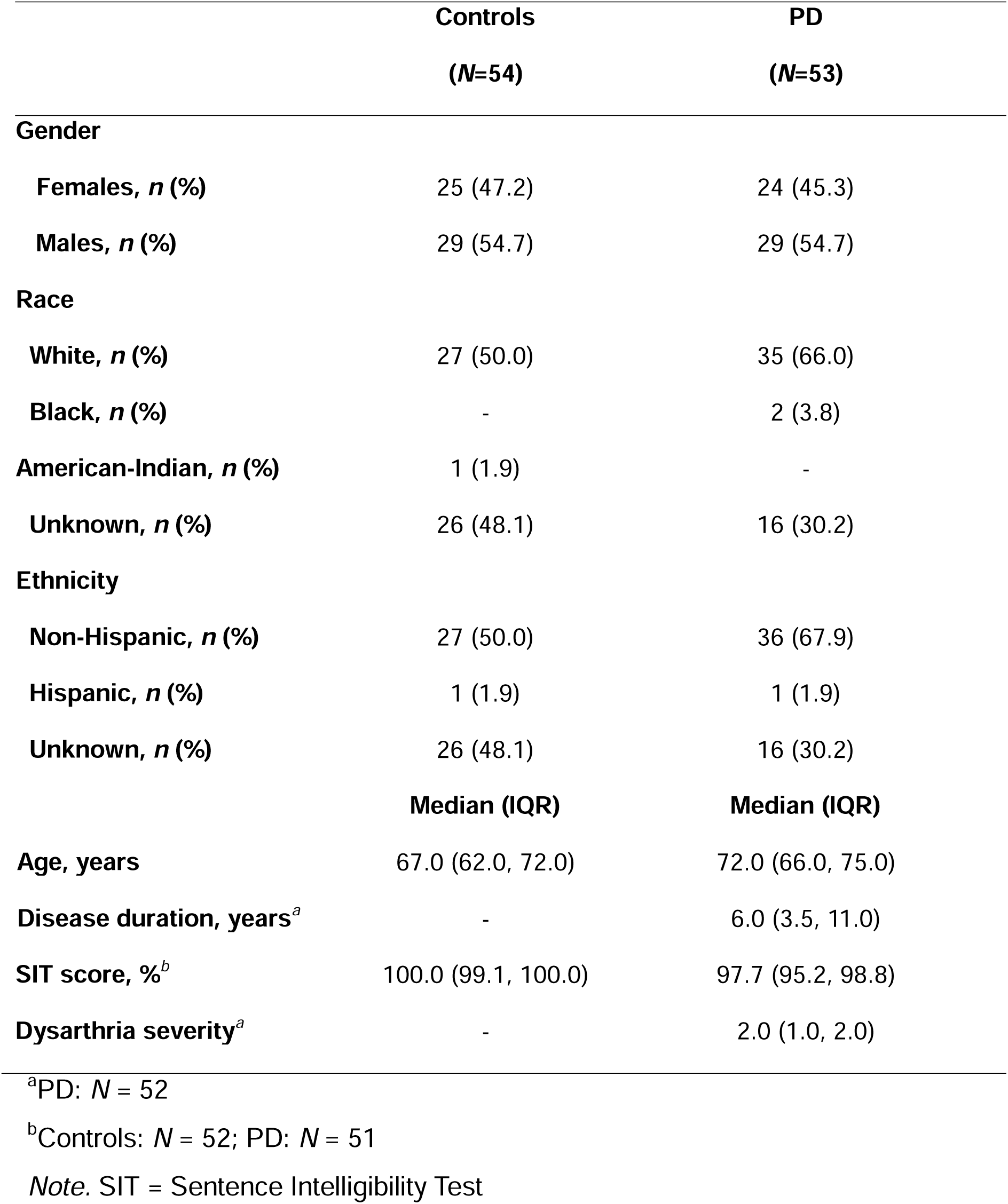
Participant Demographics.

### Kinematic Data Recording

All kinematic data recording procedures are described in detail in Mefferd et al. (2026). Briefly, all participants were asked to read the first five sentences of the Rainbow passage (Fairbanks, 1960) using their typical rate and loudness. During the reading task, articulatory movements were recorded using three-dimensional electromagnetic articulography (AG501, Carstens Medizinelektronik). Sensors were attached to the posterior tongue, tip of the tongue, midline of the upper and lower lips, as well as the jaw. The sensor of the posterior tongue was placed approximately 4cm from the tip of the tongue while the sensor on the tip of the tongue was placed approximately 1cm from the tip of the tongue. The jaw sensor was attached to the gumline in the center of the lower frontal incisors. The tongue and lip sensors were attached using a high-viscosity dental adhesive (i.e., Glustitch®) while the jaw sensor was attached using a putty (i.e., Stomahesive®). Three head reference sensors were placed on a pair of goggles, which were worn by the participant during the data collection.

The acoustic signal was recorded in a time-synchronized fashion with the kinematic data at a sampling rate of 48kHz using an omni-directional lavalier microphone. The microphone was placed approximately 15 cm away from the participant’s mouth. A large monitor was located approximately 5 feet in front of the participant and displayed the text of the reading passage for the participant.

### Kinematic Data Processing

As described in Mefferd et al. (2026), a biteplane recording was completed to transpose the kinematic data into a head-based coordinate system. During the biteplane recording, a biteplate with three additional sensors was placed between the participant’s teeth. CalcPos software (Medizintechnik Carstens) was used to convert the raw kinematic data into three-dimensional positional data. NormPos (Medizintechnik Carstens) was used to apply a head movement correction algorithm to all kinematic data and rotate the positional data into a head-based coordinate system. The resulting kinematic data for the lower lip and tongue contained contributions of the jaw. The lower lip and tongue data were not decoupled from the jaw.

As outlined in Mefferd et al. (2026), the kinematic recording of the Rainbow passage was trimmed into speech runs using SMASH (Green et al., 2013) based on onsets and offsets that were determined during an acoustic analysis of the time-synchronized speech signal in Praat (Boersma & Weenink, 2009). Specifically, speech runs were free of any pauses greater than 250ms (e.g., Allison et al., 2019) as well as non-speech noise such as coughing, laughing, or throat clearing. For each speech run, the Euclidean distance signals between the center head reference sensor (HC) and each articulator (i.e., posterior tongue, tongue tip, lower lip, jaw) were calculated in SMASH.

The trimmed kinematic speech runs were then loaded into a customized Matlab script that calculated the amplitudes of each movement segment (“stroke”). The onsets and offsets of the movement segments, or strokes, were defined by local positional minima and maxima. The amplitude of each stroke was calculated as the distance between the two positional extrema. For each articulator of each participant, stroke amplitudes were submitted to a frequency histogram analysis. However, the first and last stroke of each speech run as well as all strokes with amplitudes of less than 1mm were excluded from the frequency histogram analysis. For a more detailed description of the stroke amplitude calculation and the frequency histogram analysis please see Mefferd et al., (2026).

### Articulatory Performance Scoring Procedures

Based on the findings of the amplitude frequency distribution, each talker with PD received a score for each articulator’s amplitude performance. These scores were used to establish an amplitude performance profile for each talker with PD. The following calculations were completed: First, for each articulator a stroke frequency was calculated that included all strokes that were 6mm and larger (jaw) or 10mm and larger (lower lip, tongue tip, posterior tongue). The thresholds of 6mm (jaw) and 10mm (lower lip, tongue tip, posterior tongue) were selected based on the findings in Mefferd et al. (2026) showing a trend of talkers in the PDdys+artic subgroup producing less strokes than controls beginning at these amplitude levels (see also Figure 1). As in Mefferd et al. (2026), the large-size stroke frequency was expressed as a percentage of the total stroke count to control for the variability of the total stroke count across talkers. The resulting large-size stroke frequency was used as an indicator of hypokinesia. It should be noted that a decrease in large-size stroke frequency is mirrored by an equal increase in the small-size stroke frequency as the total stroke frequency is always 100%. In this study, we chose to score the large-size stroke frequency; however, scoring the small-size stroke frequency would also be possible and lead to the same findings.

**Figure 1.**
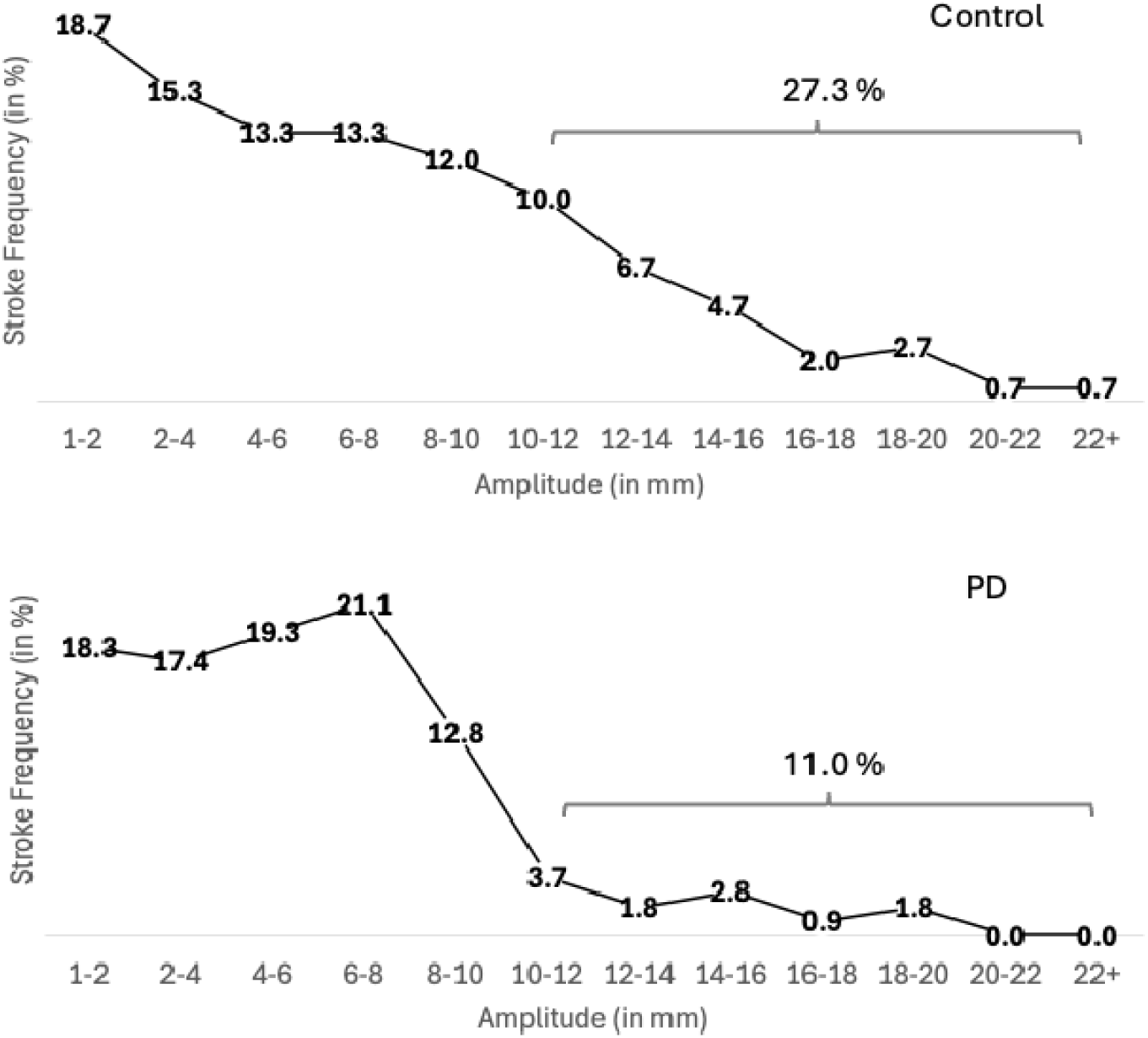
Stroke frequencies of the posterior tongue for one male control talker (top) and one male talker with PD (bottom). The bracket indicates the amplitude range used to calculate the large-size stroke frequency for each articulator of each talker

Next, the data of the control talkers’ large-sized stroke frequencies was grouped based on sex and the 12.5^th^, 25^th^, 75^th^, and 87.5^th^ percentiles of large-size stroke frequencies were calculated for female and male control talkers. These percentiles were then used to score the performance of talkers with PD based on their respective control group (female or male controls). That is, talkers with PD who had a large-sized stroke frequency that fell between the 25^th^ and 75^th^ percentile of their respective control group were deemed to have normal-sized stroke amplitudes and received a score of 0. Talkers with PD whose large-sized stroke frequencies fell between the 25^th^ and 12.5^th^ percentile were deemed to have a mild amplitude reduction and received a score of -1. Talkers with PD with large-sized stroke frequencies below the 12.5^th^ percentile were deemed to have pronounced amplitude reduction and received a score of -2. Finally, talkers with large-sized stroke frequencies between the 75^th^ and 87.5^th^ percentile were deemed to have mild amplitude exaggerations and received a score of +1 and talkers with large-sized stroke frequencies above 87.5^th^ percentile were deemed to have pronounced amplitude exaggeration received a score of +2. Figure 2 illustrates this process based on a boxplot with the landmarks of the different percentiles and the corresponding amplitude performance scores. Table 2 shows the cut-offs for each articulator and the corresponding scores.

**Figure 2.**
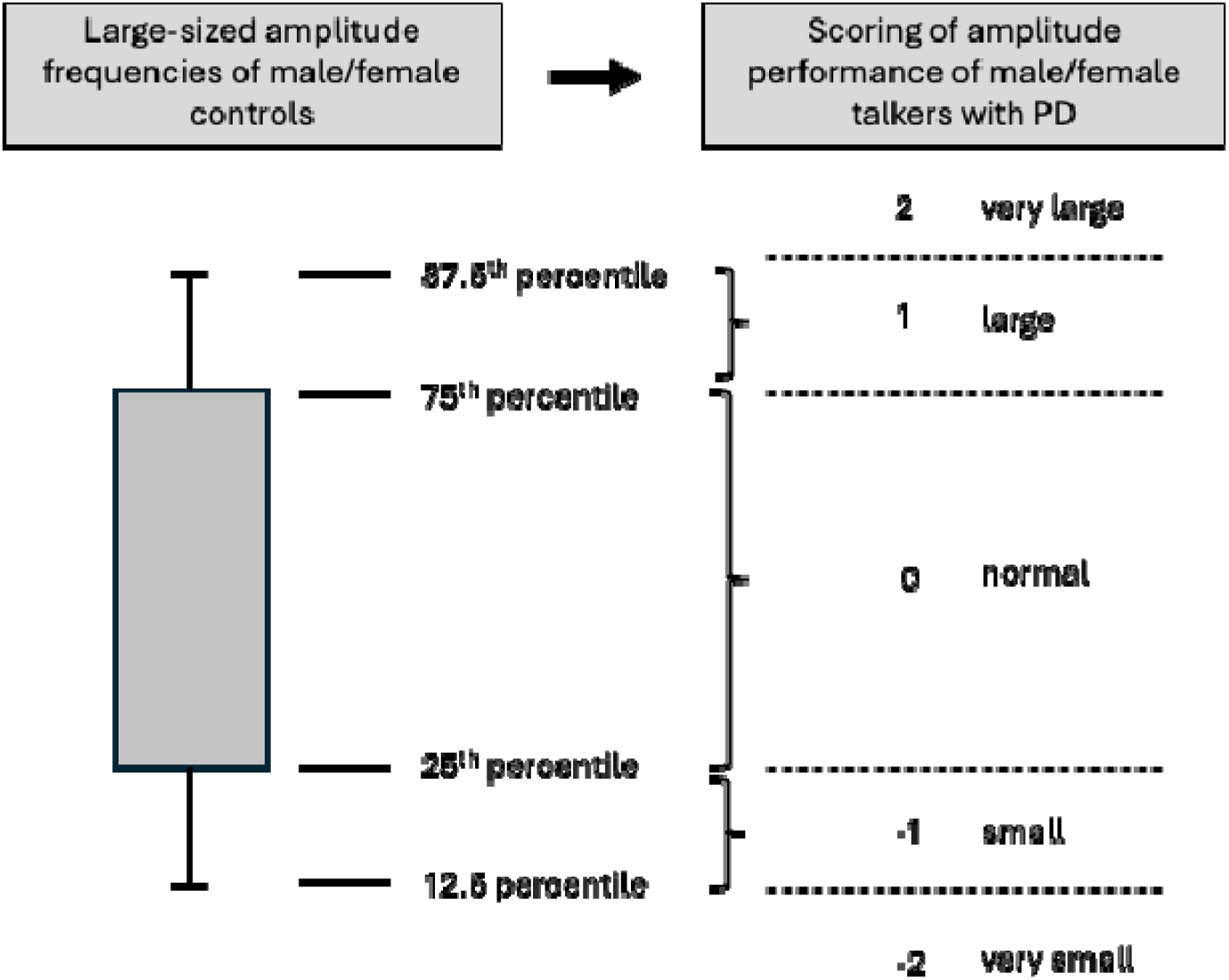
Overview of the conversion process from amplitude frequency distributions to amplitude performance scores

**Table 2.** The articulator-specific thresholds of large-sized stroke frequencies (in %) and their corresponding score for male (M) and female talkers (F)

| Percentile | Score | Jaw |  | Lower Lip |  | Tongue Tip |  | Posterior Tongue |  |
| --- | --- | --- | --- | --- | --- | --- | --- | --- | --- |
|  |  | M | F | M | F | M | F | M | F |
| 12.5 | -2 | 6.35 | 2.09 | 4.81 | 2.12 | 19.33 | 11.64 | 21.22 | 8.88 |
| 25 | -1 | 9.47 | 6.21 | 8.29 | 4.11 | 22.96 | 20.32 | 24.78 | 14.06 |
| 50 | 0 |  |  |  |  |  |  |  |  |
| 75 | +1 | 40.39 | 31.22 | 27.58 | 20.52 | 41.39 | 30.63 | 37.80 | 30.96 |
| 87.5 | +2 | 46.01 | 42.69 | 35.13 | 25.65 | 46.73 | 35.66 | 42.24 | 34.26 |

For the purpose of the current study, we consolidated the scores of the posterior tongue and the tongue tip to one overall tongue amplitude performance score to reduce the complexity and enhance the interpretability of the findings of the cluster analysis. That is, for the overall tongue score, we entered the more severe score of the two tongue scores. Table 3 provides examples of the resulting amplitude performance profiles for talkers with PD. These profiles were then used for the cluster analysis.

**Table 3.** Example of the amplitude performance profiles (cells shaded in grey) of three talkers with PD. Large-size amplitude frequencies (in %) were converted into scores.

| Talker ID | Jaw |  | Lower Lip |  | Tongue |  |
| --- | --- | --- | --- | --- | --- | --- |
|  | Frequency | Score | Frequency | Score | Frequency | Score |
| PDF32 | 8.9 | 0 | 3.9 | -1 | 16.5 (TT) | <u>-1</u> |
|  |  |  |  |  | 16.0 (PT) | 0 |
| PDF18 | 74.5 | 2 | 26.0 | 2 | 54.4 (TT) | <u>2</u> |
|  |  |  |  |  | 37.6 (PT) | 2 |
| PDF25 | 4.4 | -1 | 1.4 | -2 | 4.2 (TT) | <u>-2</u> |
|  |  |  |  |  | 2.4 (PT) | -2 |

*Note:* Frequency indicates the percent of large-size amplitudes of total amplitudes. The underlined score for the tongue indicates the overall tongue score that was submitted to the hierarchical cluster analysis.

### Demographic and Clinical Features of Talkers with PD

Demographic features of interest included the talker’s age and sex while clinical features included age at disease onset, disease duration, treatment (medication versus DBS), SIT score, dysarthria severity rating, and seven perceptual speech features of the Mayo clinic dysarthria rating scale. The procedure for the identification of the perceptual speech features is described in the following section.

### Perceptual speech features

A profile of the perceptual speech features was established for each talker with PD using the Mayo clinic dysarthria rating scale. As described in Mefferd et al. (2026), two speech language pathologists (SLPs) with clinical experience in motor speech disorders (one being the first author) rated each perceptual speech feature of the Mayo clinic dysarthria rating scale on a scale from 0 (normal) to 4 (profound) using the SIT recordings. Then, the two SLPs discussed their assessments, and established a consensus rating if discrepancies were discovered. In five cases a third SLP (the second author) was asked to weigh in to establish a consensus rating. For the purpose of the second aim of the current study, seven perceptual speech features were selected from the Mayo clinic dysarthria rating scale to test for potential differences in these features across kinematic subgroups: speech rate, increased rate in segments, short rushes of speech, imprecise consonants, reduced stress, overall loudness, and vocal decay. These features were selected due to known influences of rate and loudness manipulation on the articulatory amplitude (e.g., Mefferd & Green, 2010) and established associations between perceptible speech imprecision and articulatory range of motion (Thompson & Kim, 2025)

The severity ratings of the selected perceptual speech features (except for the feature “speech rate”) were converted to binary variables with a rating of 0 indicating the absence of the specific perceptual speech feature and 1 indicating the presence of a specific perceptual speech feature. This conversion was chosen as a first step to explore potential differences in perceptual speech features across subgroups. Specifically, collapsing severity ratings of the perceptual speech features into binary variables reduces the complexity of the statistical analysis, which was preferable given that subgroups with relatively small sample sizes were expected. Because speech rate deviated in both directions (i.e., slower and faster), the severity ratings for speech rate were retained and not converted into a binary variable.

### Statistical Analysis

The amplitude performance profile of each talker with PD consisting of three scores corresponding to the jaw, lower lip and tongue (grey-shaded cells in Table 3) was submitted to a hierarchical cluster analysis. Squared Euclidean distances were used as the measure of dissimilarity and the Ward’s method was selected to construct the cluster dendrogram to identify potential kinematic subgroups. The optimal number of clusters was determined based on the clinical relevance of the articulatory performance profiles of the talkers in the identified clusters. To determine if and how the identified subgroups differed in their amplitudes, scores of each articulator were submitted to Kruskal Wallis tests to determine between-group differences. Given that three Kruskal Wallis tests were conducted (one test per articulator), the critical alpha level was adjusted to *p* = .01. If the Kruskal Wallis test was significant, pairwise between-subgroup comparisons were conducted using Bonferroni-corrected Dunn’s tests. Non-parametric tests were chosen because per-subgroup sample sizes were expected to be relatively small and/or distributions skewed.

Due to small per-subgroup sample sizes, non-parametric tests were also chosen to determine if demographic and/or clinical features of talkers with PD were associated with specific kinematic subgroups. Because a total of 14 variables were tested, the *p*-value was adjusted to *p* = .003. Kruskal Wallis tests were completed for each clinical feature that was a continuous or ordinal variable (i.e., age, age at disease onset, disease duration, SIT score, dysarthria severity rating, rating of speech rate). Pairwise between-subgroup comparisons were conducted using Bonferroni-corrected Dunn’s tests if the Kruskal Wallis test was significant. For all binary variables (i.e., sex, perceptual speech features: presence/absence of increased rate in segment, short rushes of speech, imprecise consonants, overall reduced loudness, vocal decay, reduced stress), the Fisher-Freeman-Halton exact test was used and if significant, the minimum expected count for each cell was used to determine between group patterns.

## RESULTS

### Research Aim 1: To identify subgroups of talkers with PD based on their articulatory performance profiles using a data-driven approach

A hierarchical cluster analysis revealed six kinematic-based subgroups of talkers with PD and five talkers that did not cluster with these six subgroups. The size of the subgroups ranged from four to twelve talkers. The median and range of the scores for each of the six subgroups are displayed in Table 4. As can be seen, subgroup 2 – 7 had either median scores of 0 indicating normal amplitudes or negative median scores indicating small to very small amplitudes. Of the five talkers that did not cluster with the six kinematic subgroups, three talkers had positive scores amplitudes of either the tongue and lip, the tongue, lip, and jaw, or the jaw and lip indicating large amplitudes. The other two talkers who did not cluster with any of the six subgroups had a negative score for the jaw, a negative score or a zero score for the lower lip, and a positive score for the tongue indicating small amplitudes for the jaw, small or normal amplitudes for the lower lip, and large amplitudes for the tongue.

**Table 4.** Scores of the jaw, lower lip, and tongue amplitudes for each subgroup.

| Subgroup | *n* | Jaw |  | Lower lip |  | Tongue |  |
| --- | --- | --- | --- | --- | --- | --- | --- |
|  |  | Median | Range | Median | Range | Median | Range |
| 2 | 4 | **0** | 0-1 | **0** | - | **0** | 0-1 |
| 3 | 7 | **0** | 0-1 | **0** | 0-2 | **-1** | -1-0 |
| 4 | 10 | **0** | - | **0** | 0-1 | **-2** | - |
| 5 | 12 | **0** | -1-0 | **-2** | -2- -1 | **-2** | -2- -1 |
| 6 | 5 | **-1** | -1-0 | **-1** | -1-0 | **-1** | -1-0 |
| 7 | 10 | **-2** | - | **-2** | -2- -1 | **-2** | -2- -1 |

Kruskal-Wallis tests were conducted to determine how the subgroups differed in their articulator-specific amplitude performance scores. Subgroup 1 and the two talkers that were outliers were excluded from in these statistical analyses. If a Kruskal-Wallis test was significant, pairwise post-hoc comparisons were conducted using a Bonferroni-corrected critical alpha level of .003 (15 pairwise comparisons). Therefore, the critical alpha level was set to *p* < .003. We observed that each of the three amplitude scores differed among the six included subgroups [jaw: *H*(5) = 36.53, *p* < .001; lower lip: *H*(5) = 39.44, *p* < .001; tongue: *H*(5) = 38.68, *p* < .001]. Findings of the post-hoc pairwise comparison are provided in Table 5. Subgroup 7 had significantly lower scores for the jaw than subgroups 2 – 6 (*p* ≤ .002) indicating that talkers in subgroup 7 had significantly more pronounced amplitude reduction of the jaw compared to talkers of the other five subgroups. Post hoc pairwise comparisons showed that subgroups 5 and 7 had significantly lower scores for the lower lip than subgroups 2 - 4 indicating that talkers in subgroups 5 and 7 had significantly more pronounced amplitude reduction of the lower lip than talkers in subgroups 2 - 4 (see Table 6). Finally, subgroups 4 and 5 had significantly lower tongue scores than subgroups 2,3 and 6 (*p* ≤ .001) indicating that talkers in subgroups 4 and 5 had significantly more pronounced amplitude reduction than talkers in subgroups 2, 3, and 6 indicating that talkers in subgroups 4 and 5 had significantly more pronounced amplitude reduction than talkers in subgroups 2, 3, and 6 (see Table 7). Subgroup 7 had significantly lower tongue scores than subgroup 2 (*p* = .002) indicating that talkers in subgroup 7 had significantly more pronounced amplitude reduction than talkers in subgroup 2.

**Table 5.**
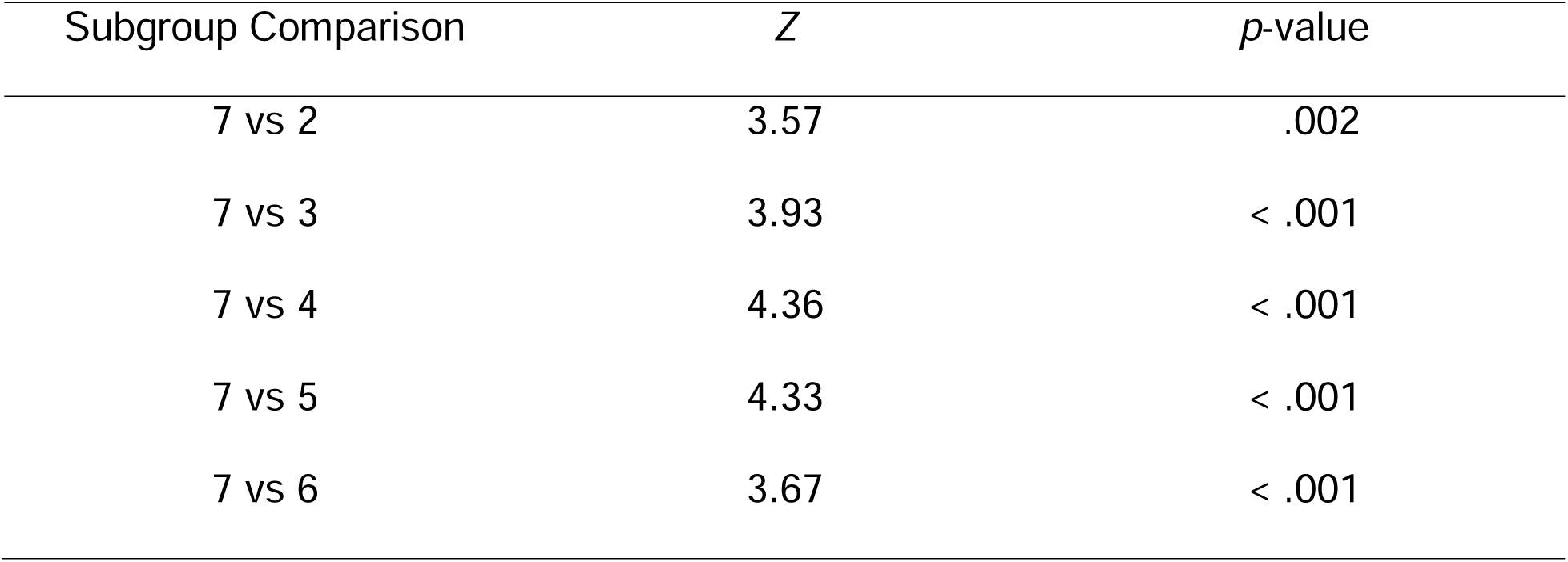
Findings of the pairwise comparison for the jaw.

**Table 7.** Findings of significant pairwise comparisons for the tongue.

| Subgroup Comparison | Z | *p*-value |
| --- | --- | --- |
| 4 vs 2 | 3.56 | .002 |
| 4 vs 3 | 3.93 | < .001 |
| 4 vs 6 | 3.69 | < .001 |
| 5 vs 2 | 3.11 | .001 |
| 5 vs 3 | 3.69 | < .001 |
| 5 vs 6 | 3.51 | .001 |
| 7 vs 2 | 3.15 | .002 |

In summary, the comparisons revealed that subgroups 4, 5, and 7 differed from subgroups 2, 3, and 6 by having significantly more pronounced amplitude reduction in one or more articulator. Subgroups 4, 5, and 7 also differed from each other in at least one articulator. While talkers in subgroup 7 demonstrated more pronounced reduction of jaw amplitudes compared to subgroups 4 and 5, talkers in subgroup 5 exhibited more pronounced reduction of lip amplitudes than subgroup 4. No significant differences in any of the three articulators were observed between subgroups 2, 3, and 6. Therefore, these three subgroups were combined and labeled subgroup 2 for subsequent analyses. Talkers in this subgroup had either normal amplitudes, mildly reduced, or mildly exaggerated movements of the jaw, lip, and/or tongue.

The small cluster of three talkers with PD detected in the cluster analysis who exaggerate their articulatory movements as well as the two talkers who exhibited amplitude reduction of the jaw and lips or only the jaw and amplitude exaggerations of the tongue were not considered to represent separate kinematic subgroups. Rather, we considered those five talkers as outliers whose performance profile did not align with any of the four main kinematic subgroups that were identified. Table 8 lists the final set of kinematic subgroups within the current cohort of talkers with PD and their amplitude performance profiles.

**Table 8.**
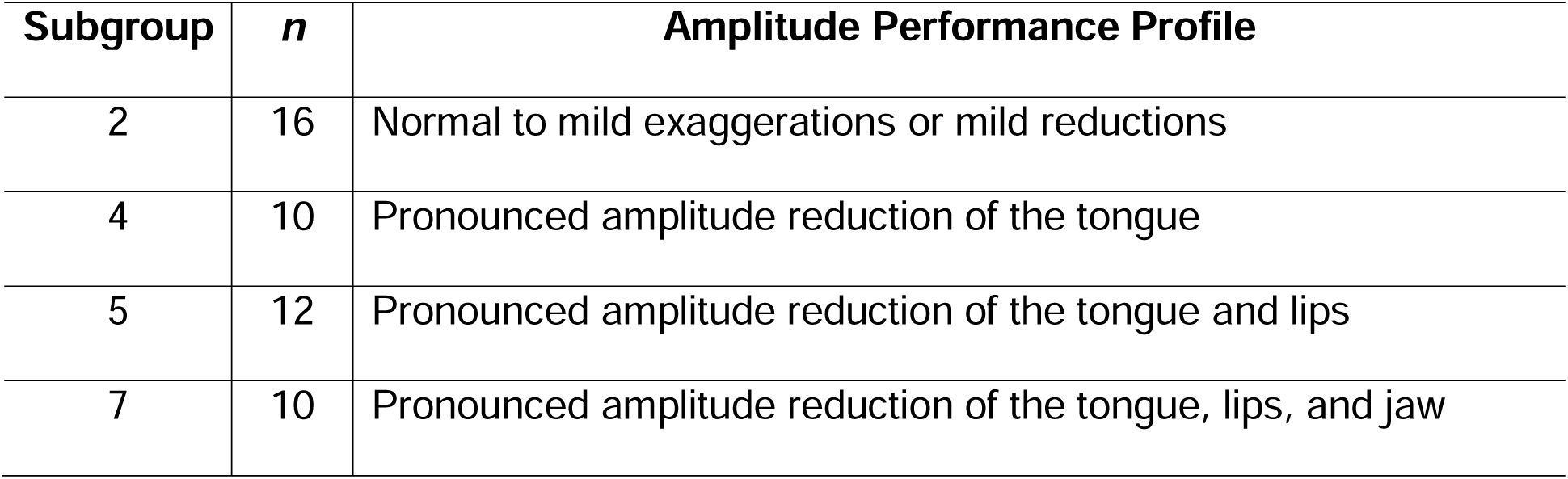
Articulatory impairment characteristics of each identified subgroup.

| **Subgroup** | ***n*** | **Amplitude Performance Profile** |
| --- | --- | --- |
| 2 | 16 | Normal to mild exaggerations or mild reductions |
| 4 | 10 | Pronounced amplitude reduction of the tongue |
| 5 | 12 | Pronounced amplitude reduction of the tongue and lips |
| 7 | 10 | Pronounced amplitude reduction of the tongue, lips, and jaw |

### Research Aim 2: To determine if kinematic-based subgroups of talkers with PD were associated with specific demographic or clinical features of talkers with dysarthria

To determine potential differences in demographic and clinical features between the four main kinematic subgroups, clinical features were submitted either to Kruskal-Wallis tests (continuous or ordinal variables) or to Fisher-Freeman-Halton exact tests (binary variables). If the Kruskal-Wallis test was significant at a *p*-value of p < .003, pairwise comparisons were conducted between the four subgroups (six between-group comparisons in total). To control for multiple comparisons during the post-hoc testing, the critical alpha level was set to *p* = .008 for the post-hoc comparisons.

Table 9 presents an overview of the statistical findings of the Kruskal-Wallis tests. Figure 3 shows the descriptive statistics of the continuous or ordinal variables for each of the four main kinematic subgroups and the two small clusters of talkers whose performance did not align with the four main kinematic subgroups. Table 10 provides an overview of the findings of the Fisher-Freeman-Halton exact tests for the binary variables. Although findings are provided for the four main kinematic subgroups and the two small clusters of talkers who do not align with the four main subgroups, the Fisher-Freeman-Halton exact tests were only based on data of the four main subgroups. As indicated in Tables 9 and 10, none of the demographic or clinical features were significantly different between the four main kinematic subgroups.

**Figure 3.**
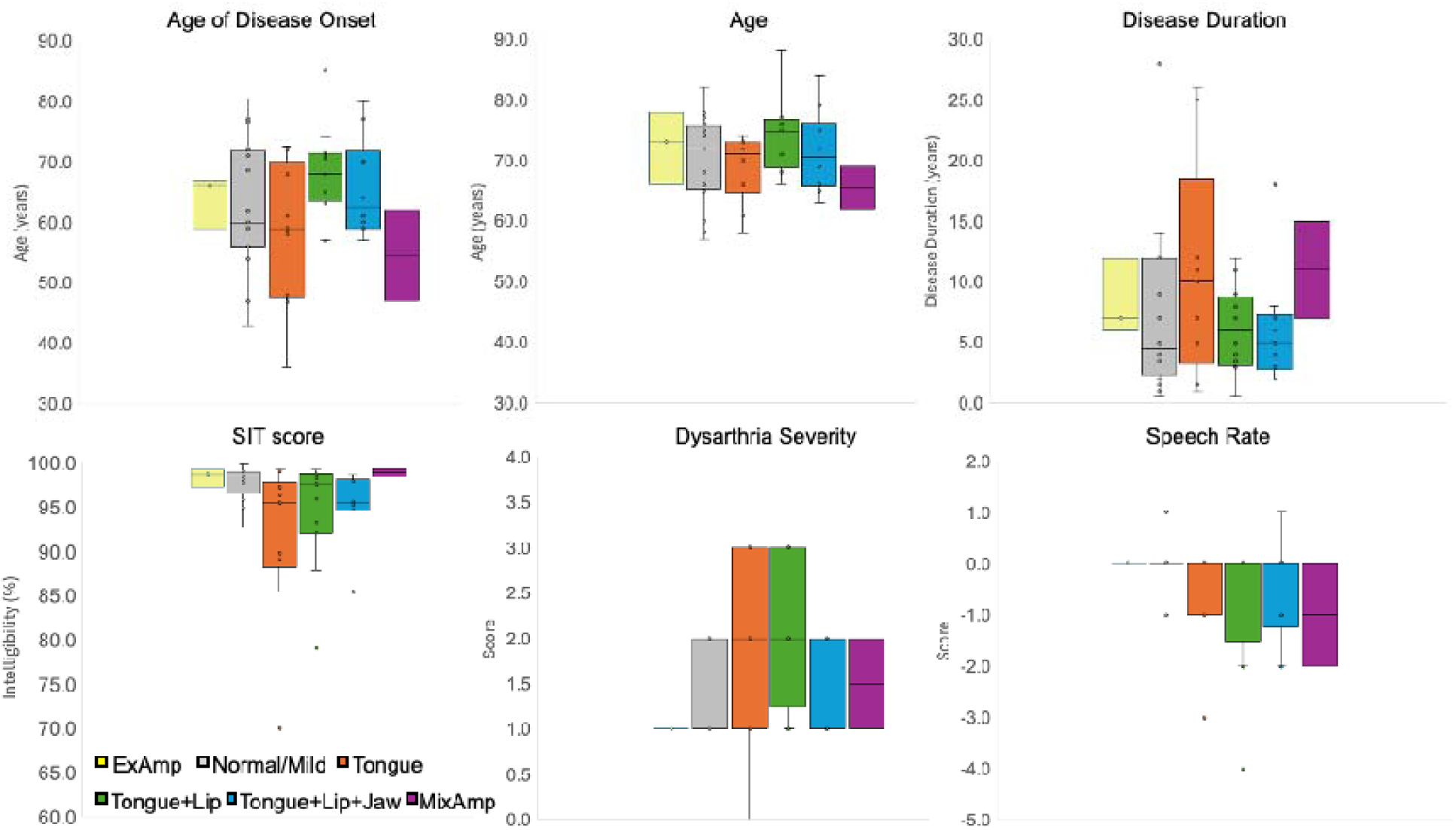
Dysarthria severity across kinematic subgroups

**Table 9.** Overview of findings of the Kruskal-Wallis tests for continuous and ordinal variables.

| Variable | Kruskal-Wallis H | *p*-value |
| --- | --- | --- |
| Age | 4.46 | .22 |
| Age at Onset | 3.96 | .27 |
| Disease Duration | 1.62 | .66 |
| SIT score | 7.04 | .07 |
| Dysarthria Severity | 7.70 | .05 |
| Speech Rate | 2.13 | .55 |

**Table 10.**
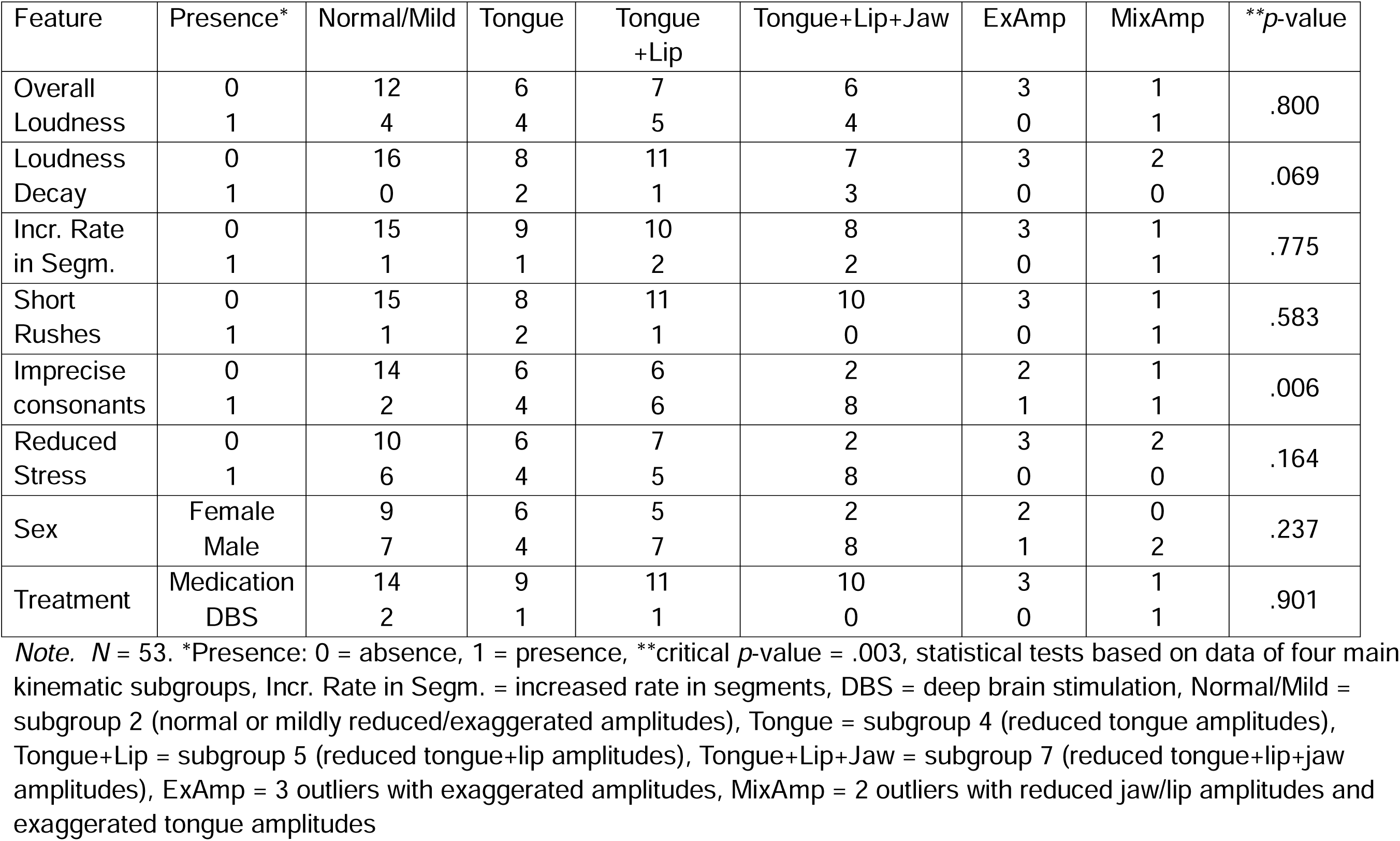
Overview of binary features across the four main kinematic subgroups and two clusters of outliers.

## DISCUSSION

This study sought to identify kinematic subgroups of talkers with PD who differ in their amplitude performance profiles. We further explored whether the identified kinematic subgroups were associated with specific demographic and/or clinical features of talkers with PD (e.g., age, sex, disease duration, dysarthria severity, specific perceptual speech features). A hierarchical cluster analysis was conducted using jaw, lip, and tongue amplitude performance scores. Initially, eight distinct kinematic-based subgroups were identified. Based on findings of the Kruskal Wallis tests, these eight subgroups were consolidated into four main kinematic subgroups. Five talkers did not align with any of the four main kinematic subgroups. One of the four main kinematic subgroups was characterized by normal, mildly exaggerated, or mildly reduced amplitudes while the other three kinematic subgroups were characterized by pronounced amplitude reduction of the tongue, lip and tongue, or jaw, lip, and tongue. The five talkers who did not align with the four main kinematic subgroups had amplitude performance profiles characterized by either exaggerated amplitudes (*n* = 3) or mixed amplitude performances (*n* = 2, reduced jaw and lip amplitudes with exaggerated tongue amplitudes). None of the demographic or clinical features yielded significant differences between the four main kinematic subgroups.

### Kinematic-Based Subgroups of Talkers with PD

The identification of the various kinematic subgroups suggests that the articulatory subsystem can be differently affected by the disease. This finding helps explain the mixed results of previous kinematic studies and suggests that discrepancies in study findings may have not merely resulted from differences in methodological approaches and experimental designs. Instead, they may have varied depending on the kinematic subgroup of talkers that was predominantly enrolled. For example, Ackermann et al. (1997) and Connor et al. (1989) reported no significant amplitude reduction of the lower lip, which aligns with the amplitude performance profiles of subgroup 2 and 4 in the current study. By contrast, Forrest et al. (1989) and Walsh and Smith (2011) reported significantly smaller lower lip amplitudes in talkers with PD compared to controls, which is congruent with the performance profile of subgroups 5 and 7 of the current study.

While all three of the main kinematic subgroups with pronounced amplitude reduction had a pronounced amplitude reduction of the tongue, only two of these three subgroups had a pronounced amplitude reduction of the lips and only one subgroup exhibited a pronounced amplitude reduction of the jaw. Thus, it appears that the tongue is in general more susceptible to hypokinesia compared to the lips and jaw. It is important to point out, however, that in the current study tongue and lower lip movements also included contributions of the jaw because the tongue and lip kinematic signals were not decoupled from the jaw. Thus, one might argue that the hypokinesia of the jaw may compound with hypokinesia of the tongue or lower lip, which may make the amplitude reduction more pronounced in the tongue and lower lip than in the jaw. However, only three of the twelve talkers with pronounced lip and tongue amplitude reduction (subgroup 5) had mildly reduced jaw amplitudes. The other nine talkers of subgroup 5 had normal jaw amplitudes. Furthermore, nine of the ten talkers with pronounced tongue amplitude reductions (subgroup 4) had normal jaw and lip amplitudes with one talker having normal jaw and mildly exaggerated lip amplitudes. Thus, it appears that hypokinesia can manifest solely in the tongue or solely in the tongue and lower lip in talkers with PD.

It is currently unclear why articulatory movements of the tongue may be particularly vulnerable to hypokinesia in talkers with PD. However, as can be seen in Table 2, jaw articulatory movements vary more in size than tongue movements in healthy controls. For example, between 9.47% and 40.39% of jaw movements produced by male control talkers had an amplitude of at least 6mm. By contrast, between 22.96% and 41.31% of tongue tip movements produced by these talkers had an amplitude of at least 12mm. Thus, tongue articulatory movements were not only larger than jaw articulatory movements, their size also did not vary as much from talker to talker as jaw amplitudes did. In other words, jaw movements can be very small in one talker and very large in another and still be considered normal whereas tongue movements must be relatively large to be considered normal.

The finding of a small number of talkers (n=3) with exaggerated amplitudes was surprising. However, this result aligns with reports of abnormally large tongue movements in studies by Wong et al. (2011) and Thies et al. (2023). The underlying mechanisms that contribute to these large amplitudes is currently unknown. However, it is possible that these talkers exaggerate their movements to maximize speech intelligibility. Perhaps these talkers were told previously that they were difficult to understand, were aware of their dysarthria, and were able to adjust their articulatory performance. We also considered the possibility that the exaggerated movements may be the result of therapeutic interventions such as the Lee Silverman Voice Treatment (LSVT) program or similar behavioral treatments since the current study did not control for a history of speech therapy. However, we could not find evidence to support this notion. Talkers who had completed a speech therapy program or were currently enrolled in one were also members of subgroups with pronounced amplitude reductions and talkers who had never been enrolled in a speech intervention program were part of the two small kinematic subgroups with amplitude exaggerations.

### Demographic and Clinical Features of Kinematic Subgroups of Talkers with PD

None of the demographic or clinical features were significantly different across the for main kinematic subgroups. The lack of significant findings may have been in part due to the relatively small sample sizes of the subgroups. However, regardless, only a few features showed trends for between-subgroup differences. One of these features was the presence of consonant imprecision, which tended to be greater in the subgroup with pronounced tongue, lip, and jaw amplitude reduction than in the subgroup with normal to mild amplitude exaggeration or reduction. Presumably, all three subgroups with pronounced amplitude reduction should have had a higher proportion of talkers with imprecise consonants than the subgroup with normal, mildly exaggerated, or mildly reduced amplitudes. However, this was not the case. Only when all three articulators were affected by hypokinesia, did most talkers have perceptible imprecision. While some of the talkers in the other two subgroups with amplitude reduction also exhibited consonant imprecision, there were also talkers who did not. It is possible that differences in the vocal tract size contributed to the discrepancies between the amplitude performance scores and the perceptual ratings for consonant precision. That is, amplitude performances that were scored as very small (pronounced amplitude reductions) may have been adequately sized for talkers with naturally smaller vocal tracts. In addition, some talkers with a pronounced amplitude reduction performed near the threshold of mild amplitude reduction, suggesting that their articulatory undershoot was not severe enough to be associated with perceived consonant imprecision. Future studies should explore alternative approaches to determine amplitude thresholds that may correspond better with clinical symptoms of dysarthria such as perceived consonant imprecision.

Interestingly, rate-related perceptual features (i.e., speech rate, increased rate in segments, short rushes of speech) were not significantly different between kinematic subgroups. There are several possible explanations for this finding. First, it should be noted that an increased articulatory rate was not a frequent perceptual feature of the talkers with PD in the current study. Although an accelerated rate and short rushes of speech are often considered hallmark speech symptoms of talkers with PD (e.g., Duffy, 2019), only twelve of the 53 talkers with PD were perceived to speak with a fast articulatory rate and only five of the 53 talkers with PD were perceived to produce short rushes of speech. Furthermore, nine of the twelve talkers with an accelerated rate exhibited amplitude reductions. However, the talkers with an accelerated rate were spread equally across the three subgroups with pronounced amplitude reduction. By contrast, the subgroup with normal, mildly exaggerated, or mildly reduced amplitudes also included two talkers with a fast speech rate. It is possible that not all talkers who were perceived to speak with an accelerated rate were objectively faster and, therefore, their rate may have not affected their articulatory amplitudes. Indeed, previous studies have shown that talkers with PD can be perceived as speaking too fast in the absence of acoustic evidence of an elevated articulatory rate (i.e., shortened segment durations, Tjaden, 2000, Weismer, 1984).

Disease duration as a clinical feature did not yield significant group differences. Nevertheless, the amplitude performance profiles of the talkers with normal, mildly exaggerated, or mildly reduced amplitudes suggest that their articulatory subsystem was less affected by the disease than that of talkers in the three subgroups with pronounced amplitude reductions. Indeed, dysarthria severity ratings tended to be lower and SIT scores tended to be higher in subgroup 2 compared to subgroups 4,5, and 7. Thus, it is possible that subgroups 2, 4, 5, and 7 reflect different stages of disease progression with talkers in subgroup 2 being at an earlier, perhaps preclinical stage, compared to subgroups 4, 5, and 7. However, it is difficult to determine if talkers in subgroups 4 and 5 are at an earlier stage than talkers in subgroup 7. Findings of previous studies suggest that articulatory impairment is first detectable in speech sounds produced by the posterior tongue and then advances to the tongue tip and lips (Logemann et al., 1978). Nevertheless, the lack of significant group differences for disease duration exemplifies how variable the onset and progression of dysarthria symptoms can be among talkers with PD. While some talkers with PD may not exhibit symptoms of dysarthria for many years after their initial diagnosis of PD, for others it can be one of the first symptoms of disease (Skodda et al., 2009). Furthermore, the lack of a significant difference in dysarthria severity or speech intelligibility between the three kinematic subgroups with pronounced amplitude reductions does not support the notion that these kinematic subgroups represent different disease stages or stages of speech decline. To better understand if PD manifest in distinctly different ways in different individuals or if the identified kinematic subgroups reflect different stages of the speech decline in PD longitudinal studies are needed.

### Clinical Implications

A better understanding of the ways PD can affect the articulatory subsystem is the first step towards a better understanding of the individual differences in the articulatory mechanisms that underlie speech intelligibility loss in talkers with PD. While the majority of talkers in the current study were more than 90% intelligible, seven talkers with PD had lower intelligibility scores ranging from 70% to 89%. Talkers with these lower SIT scores were spread across the three subgroups characterized by amplitude reduction, but they did not cluster in one specific subgroup. This finding suggests that intelligibility loss in PD may be driven by tongue amplitude reduction alone in some talkers, but may be the result of reduced lip and tongue amplitudes or reduced amplitudes of the jaw, lip, and tongue in other talkers.

It is currently unclear if talkers with different articulatory performance profiles benefit from different behavioral speech interventions. For example, it is possible that talkers with a pronounced jaw, lip, and tongue amplitude reduction may not benefit as much from a rate reduction approach (cued slow speech) as talkers with pronounced tongue amplitude reduction. That is because slow speech has been found to predominantly elicit increases in tongue amplitudes (Mefferd, 2017; Mefferd & Dietrich, 2019). By contrast, clear and loud speech cues have been shown to facilitate larger amplitudes of the tongue and jaw and, therefore, may be more effective than slow speech for talkers who exhibit jaw, lip, and tongue amplitude reductions. Future studies are warranted to empirically test these concepts.

### Study Limitations

Although the overall sample size of talkers with PD was relatively large, the identified subgroups had relatively small sample sizes, which likely contributed to the poor detections of demographic and clinical features that may be used to distinguish the kinematic-based subgroups. Future studies with larger cohorts of talkers with PD are needed to a) replicate the current findings and b) use more advanced statistical models to determine if a combination of demographic and clinical features are useful to distinguish between the various kinematic subgroups.

Another potential limitation of the current study was that tongue amplitude ratings were consolidated based on the score of the posterior tongue and the tongue tip and did not reflect a specific tongue segment. In many cases, the two segments had similar scores or one score indicated slightly less impairment than the other. Therefore, consolidating the two scores for the tongue into one score overall score was deemed as an appropriate first step. Future studies could keep both tongue segment scores for a more fine-grained analysis.

Finally, we did not decouple the jaw from the lower lip and tongue, which may have limited our ability to directly observe potential differential impairments of the jaw, lower lip, and tongue. As amplitude reduction or exaggeration of the jaw can impact the amplitude of the tongue and lower lip, the amplitude impairment of the independent tongue and lower lip movements may be under- or overestimated. However, to determine the impact of the amplitude reductions on clinical features such as dysarthria severity and specific perceptual speech characteristics (i.e., consonant imprecisions), tongue and lower lip movement could not be decoupled from the jaw.

### Conclusions and Future Directions

The findings of the current study suggest that talkers with PD vary in their amplitude performance patterns, with amplitude reductions occurring predominantly in either the tongue, the tongue and lips, or the tongue, lips, and jaw. None of the demographic or clinical features was able to distinguish the three kinematic subgroups with pronounced amplitude reduction from each other. Therefore, it may be difficult to distinguish between these three subgroups in the absence of a speech kinematic examination.

Future studies are warranted to replicate the findings of the current study. It is also important to investigate further why some talkers with PD exhibit exaggerated movements and why other talkers show reduced amplitudes of only one or two articulators while others exhibited reduced amplitudes of all articulators. Longitudinal studies are critical to determine if the three kinematic subgroups with pronounced amplitude reduction reflect different stages of the articulatory subsystem impairment or if they are distinctly different manifestations of the disease within the articulatory subsystem. If these subgroups are different manifestations of the disease, it is important to understand each subgroup’s rate of dysarthria progression and response to speech behavioral treatments. Such insights will shed light on the prognostic value and clinical significance of identifying kinematic-based subgroup.

## Data Availability

All data produced in the present study are available upon reasonable request to the first author.

## ACKNOWLEDGEMENTS

This research was funded by grant R03DC015075 and R01DC019648 from the National Institute on Deafness and Other Communication Disorders (NIDCD). Further, the recruit of participants was in part conducted via ResearchMatch, a national health volunteer registry that was created by several academic institutions and supported by the US National Institutes of Health grant UL1 TR002243 from the National Center for Advancing Translational Sciences (NCATS). The content of this manuscript is solely the responsibility of the authors and do not necessarily represent official views of NCATS or the National Institutes of Health (NIH). We would like to thank Daniel Kim, Claudia Raines, Sarah Barnes, Riley Callen, Morgan Lindstead, Nidhi Patel, Sydney Franklin, and Michael de Riesthal for their help and support with this project. Many thanks also to all study participants.

